# openEHR is FAIR-Enabling by Design

**DOI:** 10.1101/2021.02.18.21251988

**Authors:** Francesca Frexia, Cecilia Mascia, Luca Lianas, Giovanni Delussu, Alessandro Sulis, Vittorio Meloni, Mauro Del Rio, Gianluigi Zanetti

## Abstract

The FAIR Principles are a set of recommendations that aim to underpin knowledge discovery and integration by making the research outcomes *Findable, Accessible, Interoperable* and *Reusable*. These guidelines encourage the accurate recording and exchange of structured data, coupled with contextual information about their creation, expressed in domain-specific standards and machine readable formats. This paper analyses the potential support to FAIRness of the openEHR e-health standard, by theoretically assessing the compliance with each of the 15 FAIR principles of a hypothetical Clinical Data Repository (CDR) developed according to the openEHR specifications. Our study highlights how the openEHR approach, thanks to its computable semantics-oriented design, is inherently FAIR-enabling and is a promising implementation strategy for creating FAIR-compliant CDRs.

## 1. Introduction

Data semantics plays a central role in the study, design and implementation of methods and tools for extracting meaningful information from the explosion of data we have been experiencing over the past decades [1]. In particular, increasing attention has recently been dedicated to the semantic interoperability between systems and to the importance of an extensive data enrichment to improve their analysis and the preservation of their meaning when they are reused in different contexts. The crucial value of an accurate expression of semantics, both for humans and machines, is underlined also in the FAIR Guiding Principles, which strongly encourages the association of large sets of context and content metadata to the research results, to make them *Findable, Accessible, Inter-operable* and *Reusable* [2]. The FAIR Principles are general guidelines towards data-driven research and it is then up to each specific community to identify relevant domain standards and ontologies to achieve this goal. At present, the first practical implementations are mainly in the biomedical research domain [3], but the healthcare sector is also exploring the feasibility of applying the FAIR Principles [4, 5]. This paper analyses the potential FAIRness of openEHR [6], which is recognised as one the main stacks, i.e. collection of tools or standards that work together, in health informatics [7]. Starting from the openEHR specifications, we assessed the feasibility of creating a FAIR compliant resource containing Electronic Health Record (EHR) data, and showed how the openEHR design principles can handle the FAIR requirements.

## 2. Methods

### 2.1. openEHR

openEHR [6] is an open standard for e-health data, including a set of open specifications, clinical models and open-source software components, finalised to the construction of an open, vendor neutral platform for EHRs and interoperable clinical and research data.

The foundation of openEHR paradigm is the principle of separation between data representation and domain content, achieved through a multilevel information model stack. Datatypes (e.g., Text, URI), structures (e.g., single, list, tree) and containers (e.g., EHR, Composition, Demographics) are part of the *Reference Model* (RM), while the domain-related content is expressed through archetypes and templates in the *Archetype Model* (AM). Only the RM structures are implemented in software, thus decoupling the deployment of systems from the heterogeneity and variability of data. Archetypes are used to model specific domain concepts with all the possible data points that would be universally needed in any context. Technically, archetypes are extensible formal constraint definitions of object structures expressed in the Archetype Definition Language (ADL), where each data point corresponds to a specific path and can be bound to external terminologies and ontologies, to better specify its meaning. Built for specific use cases, templates define a tree of one or more archetypes, possibly hiding non-relevant data points and tightening existing constraints while maintaining all the terminological binds. Templates are used at runtime to create forms and data instances as well as to validate data input, ensuring that all data conform to the constraints defined in the included archetypes or in the template when tighter. Notably, templates preserve the path structure of each archetype’s node, ensuring in this way the inclusion in the data instances of the notion of which archetypes were used at data creation time. Paths are expressed in an XPath-compatible syntax and contain archetype attribute names and node identifiers, therefore supporting semantic querying. Queries are defined in AQL (Archetype Query Language), combining SQL and paths drawn from the archetypes.

openEHR clinical models are computable artifacts to model healthcare contents, consisting of archetypes which can be created from scratch or reusing previously published models. The openEHR Clinical Modelling Program gathers a wide international community of clinicians, researchers and developers working on the design and maintenance of the clinical models, collected in the international Clinical Knowledge Manager (CKM). At present, it includes a library of 535 governed archetypes (i.e., over 8000 data points) and other local CKMs exist too, to address specific projects’ or national needs.

### 2.2. Analysis Approach

The hypothesis of the innate FAIR compliance of the openEHR approach is primarily justified by the possibility of establishing a direct mapping between the openEHR foundational principles and the FAIR dimensions. In the archetype-driven openEHR architecture, in fact, archetypes and templates perform two key functions: (i) *semantics preservation*, long term and implementation independent, by defining structured and detailed open models for data validation at data capture or import time, therefore supporting the “I” and “R” dimensions; (ii) *semantic querying*, reachability of every data item through “semantic paths” deriving from the composition of archetypes within a template, mechanism relevant for the “F” and “A” dimensions.

In order to check this hypothesis, we have analysed if and how it would be possible to create an openEHR-based Clinical Data Repository (CDR) fulfilling the requirements expressed by each specific FAIR Principle. In particular, in this analysis we considered as “data” all the instances of clinical information in the repository EHR, and we defined two main categories of “metadata”: (i) *instance metadata*, that are all the relevant attributes describing the data, included at various levels in the EHR data instances, further sortable into *general* (such as authorship, copyright, licences, languages and translations) and *contextual* (attributes describing information on data acquisition, such as protocol and subject’s state); (ii) *knowledge metadata*, the archetypes and templates representing the domain-content models associated with the data. The inclusion of the clinical models in the metadata is motivated by the fact that they are detailed and structured sets of attributes describing the real world contents which generated the data, since the creation and validation of openEHR data instances is controlled by the knowledge artefacts derived from these specific models.

## 3. Results

Table 1 illustrates the results of our theoretical FAIRness analysis for a generic EHR system, implemented following the openEHR specifications and evaluated from the perspective of the compliance with each single FAIR Principle. As shown in detail in the table, openEHR envisages by design all of the structural components necessary to create a FAIR resource, since every FAIR Principle can be fulfilled by developing a Clinical Data Repository according to the openEHR design fundamentals and specifications.

**Table 1.**
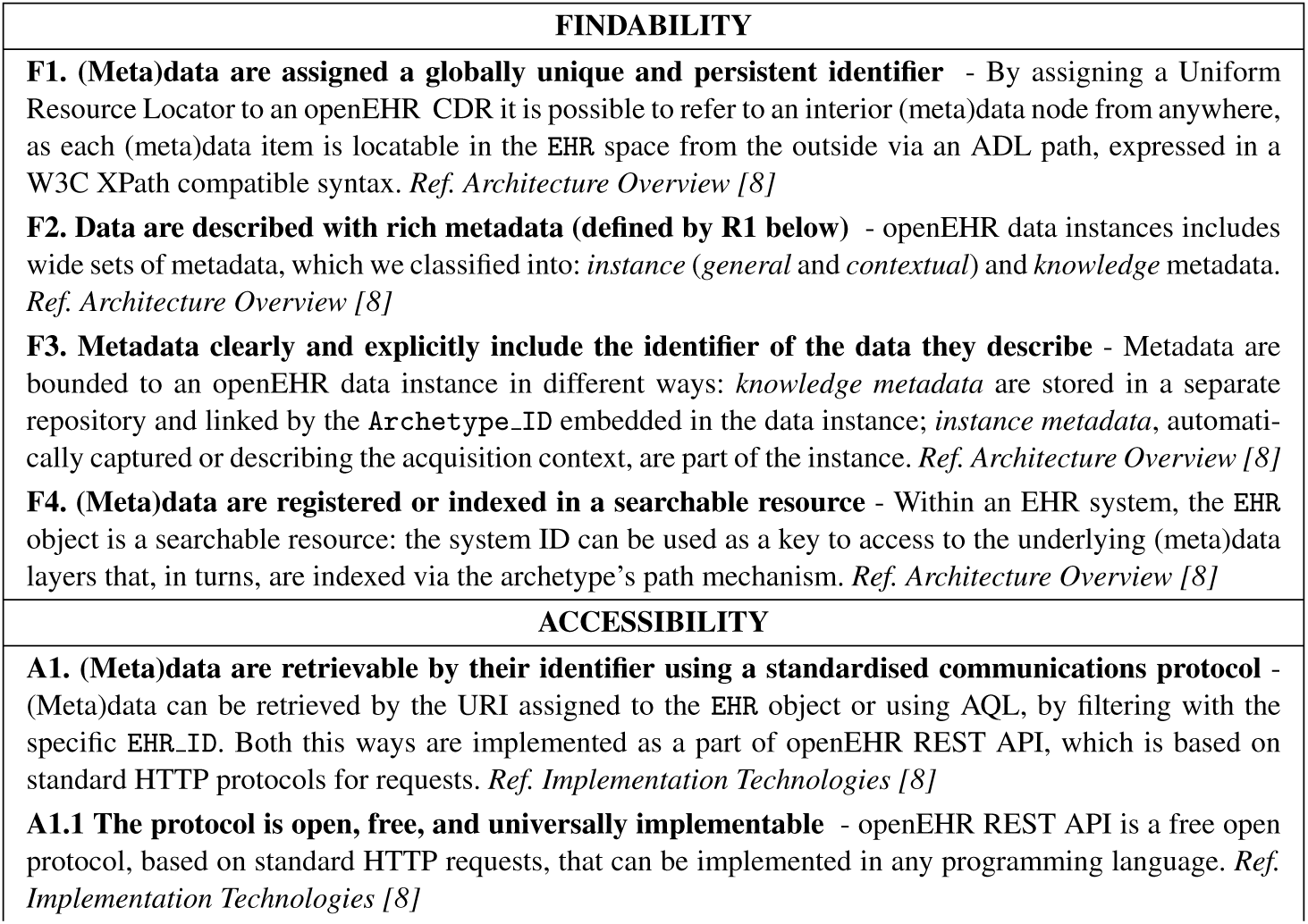
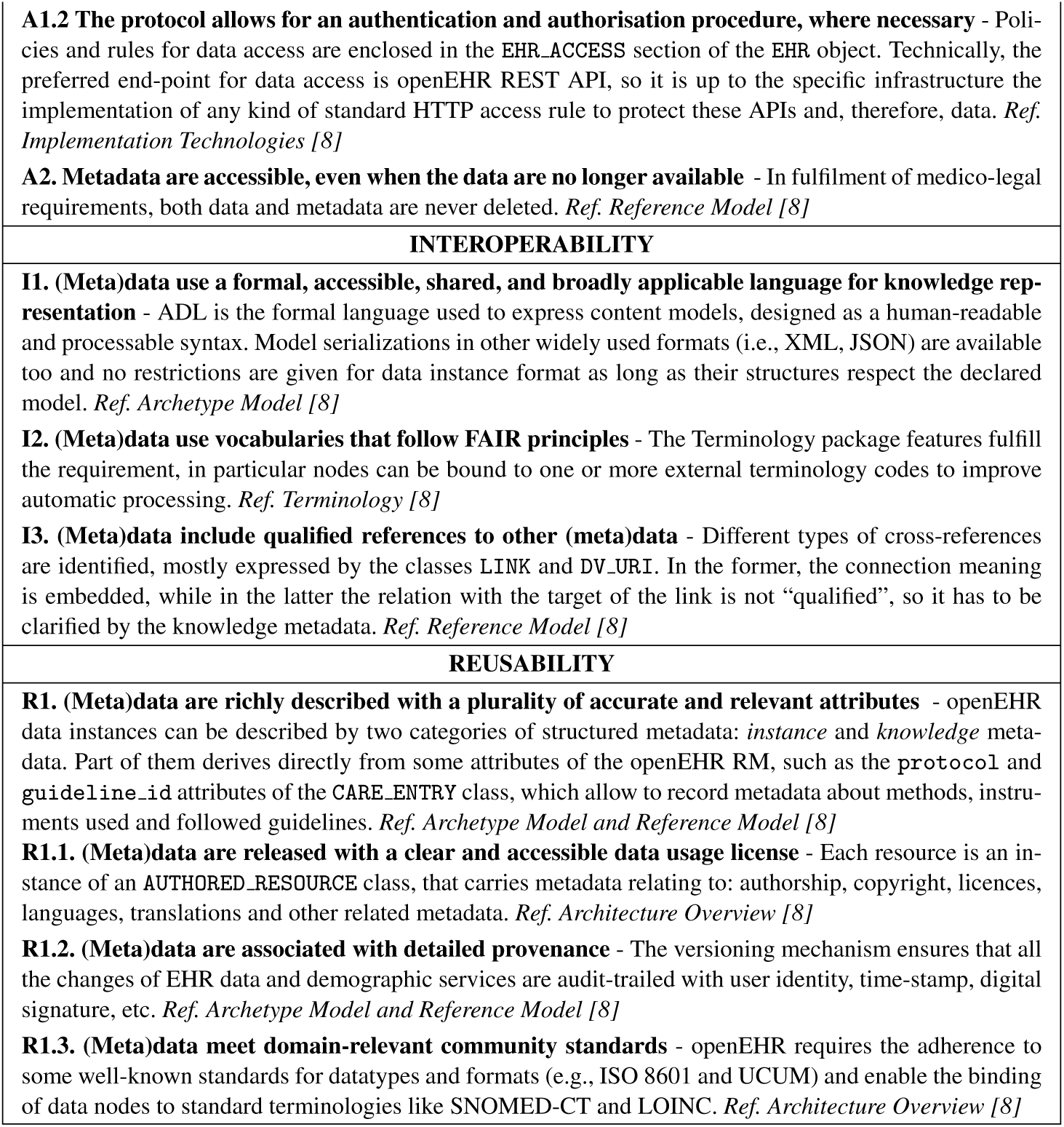
Compliance of an openEHR-based Clinical Data Repository with the FAIR Principles

## 4. Discussion

This paper shows how openEHR design fundamentals can comply with the FAIR Principles and, to the best of our knowledge, this is the first study highlighting this conceptual analogy. Only a previous work, in fact, already explored the FAIRness of a resource containing openEHR archetypes and templates [9], but its focus was on the fulfillment of the Principles for a determined repository rather than on the overall affinity between openEHR and FAIR.

Our objective was to examine the interrelation between the basis of two sets of theoretical guidelines, not the evaluation of a specific implementation. We therefore have highlighted how openEHR specifications natively support the creation of a FAIR data resource, not that any openEHR repository is FAIR. Indeed, the effective FAIRness of a repository also depends on the construction of the persistence solution and it has to be assessed for any repository, whatever kind of data it contains.

A central point of the study is the very specific assumption about the definition we adopted for metadata, specialised to include the openEHR clinical models. However, our wider interpretation is consistent both with the general FAIR suggestions about metadata [10] and with other relevant guidelines for metadata formalisation, like those enounced by the Research Data Alliance. Therefore, we can conclude that even if the assumption is very tailored to the openEHR context, it doesn’t affect the reliability of the results. Moreover, this approach enforces Interoperability and Reusability, explicitly supporting the availability of detailed description of domain knowledge.

## 5. Conclusion

Our analysis reveals the intrinsic potential of openEHR to build a FAIR-compliant Clinical Data Resource, by applying the openEHR specifications in combination with some *ad hoc* deployment configurations. This native support to FAIRness is a direct consequence of the centrality of semantic preservation and querying in the openEHR philosophy since its inception, long before the FAIR Guiding Principles were formulated. Despite its abstract nature, our analysis highlights how the openEHR approach can be considered a viable choice to have data more Findable, Accessible, Interoperable and Reusable in the clinical and biomedical context. Future work will involve exploring how this considerations can be applied to real implementations, by assessing the actual FAIRness of openEHR resources developed for a particular use case.

## Data Availability

In the manuscript there are no data.

## 6. Acknowledgments

We dedicate this work to the memory of Gianluigi Zanetti, whose legacy continues to inspire us. This work has been partially supported by the DIFRA Project (funded by the Sardinian Regional Authority) and by the European Joint Programme on Rare Diseases (grant agreement N. 825575).

